# Alzheimer’s Disease biological PET staging using plasma p217+tau

**DOI:** 10.1101/2024.01.11.24301180

**Authors:** Azadeh Feizpour, Vincent Doré, Natasha Krishnadas, Pierrick Bourgeat, James D. Doecke, Ziad S. Saad, Gallen Triana-Baltzer, Simon M. Laws, Rosita Shishegar, Kun Huang, Christopher Fowler, Larry Ward, Colin L. Masters, Jurgen Fripp, Hartmuth C. Kolb, Victor L. Villemagne, Christopher C. Rowe

**Affiliations:** The Florey Institute of Neuroscience and Mental Health, The University of Melbourne, Parkville, Victoria, Australia; Department of Molecular Imaging & Therapy, Austin Health, Melbourne, Victoria, Australia; The Australian e-Health Research Centre, CSIRO, Melbourne, Victoria, Australia; The Australian e-Health Research Centre, CSIRO, Brisbane, Queensland, Australia; Centre for Precision Health, Edith Cowan University, Joondalup, Western Australia, Australia; Neuroscience Biomarkers, Janssen Research and Development, La Jolla, CA, USA; Collaborative Genomics and Translation Group, Edith Cowan University, Joondalup, Western Australia, Australia; Curtin Medical School, Curtin University, Bentley, Western Australia, Australia; School of Psychological Sciences and Turner Institute for Brain and Mental Health, Monash University, Melbourne, Victoria, Australia; Department of Psychiatry, University of Pittsburgh, Pittsburgh, PA, USA; Florey Department of Neuroscience and Mental Health, The University of Melbourne, Melbourne, Victoria, Australia

## Abstract

**Background:** Plasma phospho-tau biomarkers, such as p217+tau, excel at identifying Alzheimer’s Disease (AD) neuropathology. However, questions remain regarding their capacity to inform AD biological PET stages at group level and maintain the same precision at individual patient level.

**Method:** Participants included 248 cognitively unimpaired (CU) and 227 cognitively impaired (CI) individuals, with Janssen plasma p217+tau Simoa® assay, ^18^F-NAV4694 Aβ PET (A) and ^18^F-MK6240 tau PET (T) data. Biological PET stages were defined based on the draft NIA-AA Revised Criteria (July 2023): Initial (A+T-), Early (A+T_MTL_+), Intermediate (A+T_MOD_+), and Advanced (A+T_HIGH_+). We used thresholds for A+ of 25 Centiloid and for T_HIGH_ of 80 Centaur (2.68 SUVR_temporo-parietal_). Adding an A-T- stage for comparison, we assessed the performance of p217+tau in discriminating between these stages at the group level using Receiver Operating Characteristic (ROC) analysis and at the individual level using logistic regression.

**Results:** Plasma p217+tau concentrations increased across the stages, with significant differences between them, except for the Initial and Early stages. Screening for Advanced (*vs*. lower stages), combined Intermediate/Advanced (*vs*. lower stages), or all stages (*vs*. A-T-), p217+tau showed good group-level discriminations (AUC 0.91, 0.92 and 0.92; CI only: AUC 0.83, 0.89, 0.93, respectively). At the individual level, the likelihood of PET stage *vs*. p217+tau level showed good discrimination of A-T- vs any A+ stage and of combined Intermediate/Advanced disease stage vs lower stages in the CI.

**Conclusion:** In addition to accurately screening for A+ individuals, plasma p217+tau shows promise for separating persons with either Intermediate or Advanced stage AD from those at a lower stage, providing prognostic information and informing better selection for trials and disease modifying therapies.

## Introduction

The *in vivo* staging of Alzheimer’s disease (AD) has become increasingly important, particularly in light of the recent breakthroughs in disease-modifying therapies for AD ^1^. Positron Emission Tomography (PET) imaging allows *in vivo* staging however, the cost and limited scalability underscores the need to assess plasma biomarkers for this purpose. Among a range of plasma biomarkers, measures of phosphorylated tau (p-tau) at amino acid 217 (p-tau217) have demonstrated significant potential, rivalling the gold-standards of amyloid-beta (Ab) PET and cerebrospinal fluid (CSF) ^2^.

The plasma “p217+tau” assay is a high sensitivity Simoa assay using a capture antibody (pT3), raised against tau paired helical filaments (PHF), that binds to phosphorylation at aa217 (p217) with enhanced binding when other nearby phosphorylated sites are present, predominantly at aa212. It has shown a similar performance to p-tau217 in detecting Aβ status in CSF, distinguishing clinical diagnostic groups and predicting progression from Mild Cognitive Impairment (MCI) to AD ^3^. A comparison of its performance to recent-generation PET tracers has demonstrated similarly good diagnostic accuracy ^4^. Furthermore, we have previously shown that p217+tau predicted cognitive decline and if used as a pre-screening tool in pre-clinical AD trials or screening tool in MCI/AD trials, substantial cost reduction could be achieved ^5^. While accumulating evidence underscores the excellent diagnostic, prognostic, and clinical trial utility of plasma p-tau biomarkers, their performance in disease staging is at present understudied. Disease staging needs to be assessed at both group level and individual level. Group-level analysis establishes receiver operating characteristic (ROC)-based thresholds (such as 90% sensitivity or 90% positive predictive value (PPV) thresholds) that have proven useful in enriching clinical trial populations. Individual-level analysis provides the foundation for development of risk assessment models aimed at the identification of individuals at elevated risk for specific stages of AD to tailor their prognosis and treatment.

A recent report by Jack *et al.* ^6^ examined whether p-tau217 could discriminate between AD disease stages. They defined four tau PET topographic stages using Braak-like PET staging schemes ^7, 8^ and group-level results indicated a good discrimination between Braak 0 and Braak 1-6 stages in the cognitively unimpaired participants.

In the present study, we were interested in both individual-level and group-level discrimination of disease stages. Therefore, we employed the biological PET staging of AD, recently proposed by draft NIA-AA Revised Diagnosis and Staging Criteria (2023) ^9^ and aimed to assess the ability of plasma p217+tau to discriminate individuals that fall within these stages.

## Methods

### Participants

From AIBL and ADNeT cohorts, 475 participants with ^18^F-MK6240 tau PET, ^18^F-NAV4694 Aβ PET and plasma p217+tau were selected. Details of cohort recruitment and evaluation are described elsewhere ^10^. Briefly, all participants were clinically classified as cognitively unimpaired (CU), or participants with mild cognitive impairment (MCI), Alzheimer’s disease (AD) dementia or non-AD dementia, by a multi-disciplinary panel blind to PET imaging and blood assays results. The MCI and dementia groups were combined into a cognitively impaired (CI) group for some analyses. A diagnosis of CU required performance within 1.5 standard deviations (SD) of the published norms for their age group on selected neuropsychological assessments. A diagnosis of MCI or AD dementia followed the internationally agreed clinical criteria ^11, 12^. Approval was obtained from institutional ethics review committees for the AIBL and ADNeT studies and written informed consent was obtained from all participants.

### PET Image Acquisition and Analysis

Aβ PET imaging involved a 20-minute acquisition, performed 50 minutes after injection of 200 MBq of ^18^F-NAV4694 intravenously. Tau PET imaging was conducted on a separate day, with a 20-minute acquisition, performed 90 minutes after intravenous administration of 185 MBq of ^18^F-MK6240. For Aβ PET scans, spatial normalization was achieved using CapAIBL ^13^ and the results were standardised using the Centiloid (CL) scale ^14, 15^. For tau PET scans, spatial normalization was performed using the MR-less CapAIBL PCA-based method ^16^. Tau PET scans were scaled using the cerebellar cortex as the reference region. ^18^F-MK6240 standardized uptake value ratio (SUVR) was estimated for three in-house composite regions of interest (ROI): 1) mesial temporal (Me) ROI, comprising entorhinal cortex, amygdala, hippocampus, and parahippocampus, 2) temporoparietal (Te) ROI, consisting of inferior temporal, fusiform, supramarginal and angular gyri, posterior cingulate/ precuneus, superior and inferior parietal, and lateral occipital cortex and 3) rest of neocortex (R) including dorsolateral and ventrolateral prefrontal, orbitofrontal, gyrus rectus, superior and middle temporal, and anterior cingulate ^17^. A CL threshold of 25 was selected to discriminate Aβ positive (A+) *vs*. Aβ negative (A-) PET scans. Previously reported thresholds based on the 95^th^ percentile of A-CU were used to discriminate tau positive (T+) from tau negative (T-) PET scans—1.18 SUVR for Me, 1.24 SUVR for Te and 1.08 SUVR for R ^18^. For tau PET uptake in the moderate SUVR range in the Te ROI, we considered values between 1.24 and 2.68. The upper threshold of 2.68 Te SUVR corresponds to 80 CenTauR ^19, 20^. We considered 80 CenTauR as a cut-off point between low/medium (or intermediate) and high tau burden, as we calculated that this approximates the threshold used for tau PET staging (intermediate *vs*. high) in the Trailblazer-Alz 2 randomized clinical trial ^1^.

### Plasma p217+tau Assay

Fasting blood was collected 2.4 ± 10.4 months from the time of Aβ PET scan and 0.1 ± 4.2 months from the tau PET scan. Plasma from K2-EDTA tubes (7.5 mL S-monovette 01.1605.008, Sarstedt), containing prostaglandin E1 (33 ng/mL of whole blood, Sapphire Biosciences), was centrifuged at room temperature for 10 mins at 200g to collect platelet-rich plasma. Then, to provide plasma that was snap frozen within 2 hr of collection, it was centrifuged at 800g for 10 mins, following which it was stored in vapour phase liquid nitrogen prior to shipping on dry ice from Australia to Janssen R&D, La Jolla, CA, USA. The plasma p217+tau assay was completed on a Single Molecule Array (Simoa®) HD-X platform, blinded to all subject data. The technique has been previously described by Triana-Baltzer *et al*. ^21^. Plasma p217+tau assay results from two laboratories, Janssen R&D (cohort 1; n=397) and Quanterix Corp, MA, USA (cohort 2; n=317), were combined. This study included all participants from cohort 2, and those unique to cohort 1 who were not already part of cohort 2 (final n=506). Assay scales measured from both laboratories were comparable.

### Implementation of 2023 Proposed NIA-AA Staging of AD with PET

In the draft NIA-AA Revised Diagnostic and Staging Criteria (2023) ^9^, the following biological PET staging scheme is proposed: 1. Initial stage: proposed as abnormal Aβ with no tau uptake (A+T-), 2. Early stage: as abnormal Aβ with tau uptake limited to medial temporal region (A+T_MTL_+), 3. Intermediate stage: as abnormal Aβ with moderate tau uptake in a neocortical ROI (A+T_MOD_+), and 4. Advanced stage: as abnormal Aβ with high tau uptake in the same neocortical region (A+T_HIGH_+). For stages 3 and 4, we considered Te as our neocortical ROI. Table 1 displays our selected CL and SUVR cut-offs and ranges for defining the aforementioned stages:

**Table 1.**
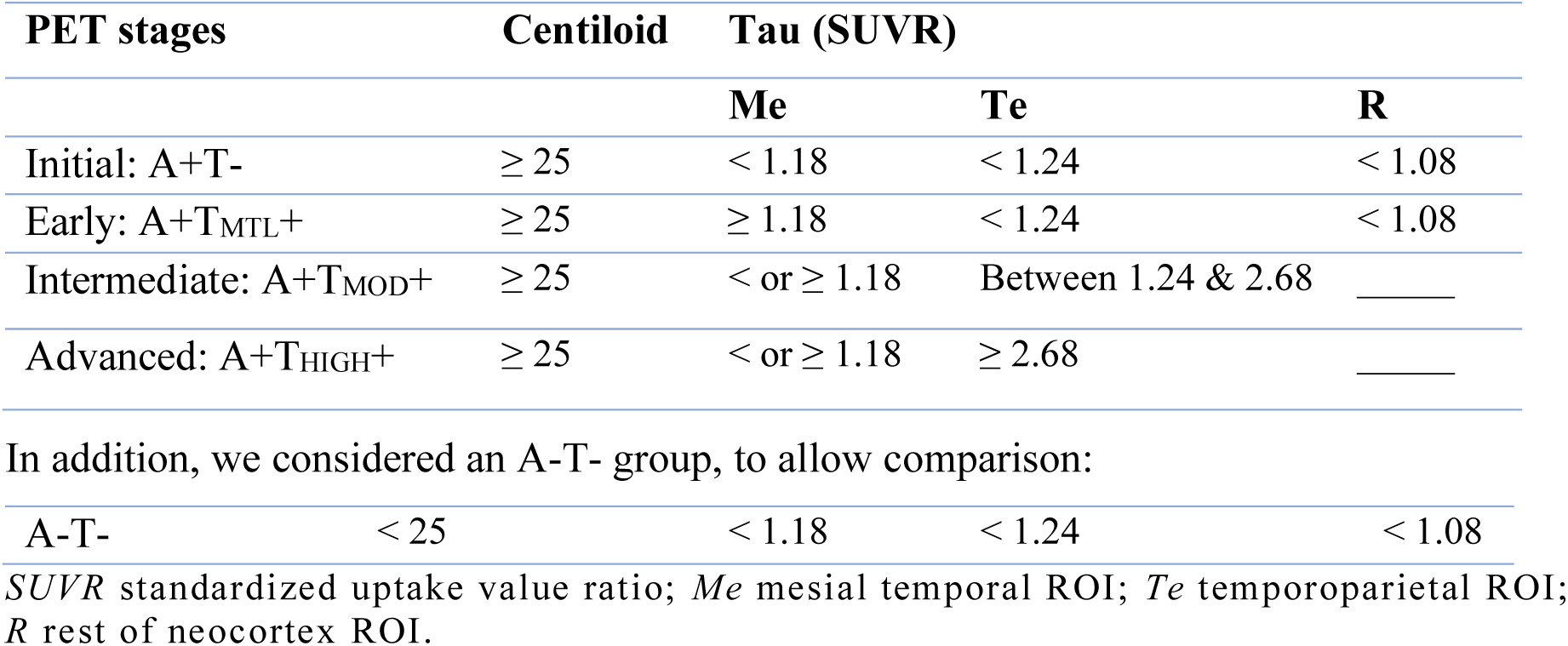
Biological PET staging.

| PET stages | Centiloid | Tau (SUVR) |  |  |
| --- | --- | --- | --- | --- |
|  |  | Me | Te | R |
| Initial: A+T- | ≥ 25 | < 1.18 | < 1.24 | < 1.08 |
| Early: A+T<sub>MTL</sub>+ | ≥ 25 | ≥ 1.18 | < 1.24 | < 1.08 |
| Intermediate: A+T<sub>MOD</sub>+ | ≥ 25 | < or ≥ 1.18 | Between 1.24 & 2.68 | _____ |
| Advanced: A+T<sub>HIGH</sub>+ | ≥ 25 | < or ≥ 1.18 | ≥ 2.68 | _____ |
| In addition, we considered an A-T- group, to allow comparison: |  |  |  |  |
| A-T- | < 25 | < 1.18 | < 1.24 | < 1.08 |

*SUVR* standardized uptake value ratio; *Me* mesial temporal ROI; *Te* temporoparietal ROI; *R* rest of neocortex ROI.

Of the 506 participants with available plasma p217+tau assay results, 31 who were A- but T+ in one or all three ROIs did not fit within any of the above AT categories and were excluded from subsequent analyses. Among these, 11 had mildly elevated MTL binding and were postulated to have primary age related tauopathy (PART) but 9 were positive on quantification in MTL and temporo-parietal. This proportion of the total study sample of ∼2% as having clearly positive neocortical tau PET but negative amyloid PET is consistent with our previous report ^22^ (see Supplementary Table 1 for demographic characteristics of the excluded participants).

### Statistical Methods

All statistical analyses were completed using Python version 3.9.13, unless otherwise specified. Demographic differences between groups were assessed using one-way ANOVA for continuous data and Pearson’s χ2 test of independence for categorical variables (gender, Apolipoprotein E (*APOE*) ε4 status, clinical diagnosis). Pairwise comparisons for continuous data were performed using Tukey Honest Significant Difference (HSD) test while categorical variables were analysed with Pearson’s χ2 test. Bonferroni correction was applied to all multiple pairwise comparisons. To investigate the magnitude of differences in plasma p217+tau concentration across the PET stages, effect sizes were estimated using Cohen’s *d*. ROC analysis was used to assess group-level discriminatory capability. The area under the ROC curve (AUC) from each comparison was used to assess the performance of p217+tau alone or in combination with other predictors (age, gender and *APOE* ε4 status) in discriminating different PET stages. The AUCs were compared using DeLong test (R version 4.2.3). Specificity, sensitivity, PPV and negative predictive value (NPV) were reported with bootstrapped 95% confidence intervals (shown in square brackets). PPV and NPV were not adjusted for disease prevalence. Optimal p217+tau thresholds for discriminating between the biological PET stages were derived using Youden’s Index.

Given that there were only nine participants with p217+tau values above 500 fg/ml, with a range from 507 to 902 fg/ml (6 of whom were A+T_HIGH_+ and 3 were A+T_MOD_+) and considering the limited statistical power to estimate the line of best fit, we restricted our regression analyses to plasma p217+tau concentrations between 0 and 500 fg/ml in the subsequent sections. To evaluate the probability of a participant belonging to a distinct PET stage (individual patient-level staging), we employed two methods. In the first approach, we computed probability scores by applying a binary logistic regression model. In this model, we used plasma p217+tau as the predictor (independent variable) and grouped PET stages as the outcome (dependent variable), where the stages were categorized into binary groups of 0 and 1. Then, we compared the results of the logistic regression to the second approach, a sliding window method. Here, we calculated PET stage probabilities within sequential bands of plasma p217+tau concentrations, with each band spanning 80 fg/ml. We selected an 80 fg/ml band width as a balance between achieving overly smoothed curves and avoiding excessive noise. The calculated probability for each band was centred at the midpoint of the p217+tau concentration range. The probability estimates from both methods were plotted against p217+tau concentrations and compared across the biological PET stages.

## Results

### Participant demographics

Demographic characteristics of the participants are presented in Table 2. Participants included 248 CU, 144 with MCI and 83 with dementia (total n = 475). Among these, 192 were categorized as A-T-, 80 as A+T-, 31 as A+T_MTL_+, 128 as A+T_MOD_+, and 44 as A+T_HIGH_+. The A+T_HIGH_+ group had a younger average age and lower level of education compared to the other groups. No significant gender differences were observed among the groups. Relative to A-T-, all other groups displayed higher rates of *APOE* ε4 carriership and greater prevalence of MCI or dementia diagnosis. These groups also exhibited higher Centiloid values and plasma p217+tau concentrations. The A+T_MOD_+ and A+T_HIGH_+ had lower MMSE scores, and A+T-, A+T_MOD_+, and A+T_HIGH_+ had higher CDR-SoB scores. For a breakdown of participant demographics by clinical diagnosis, see Supplementary Table 2.

**Table 2.**
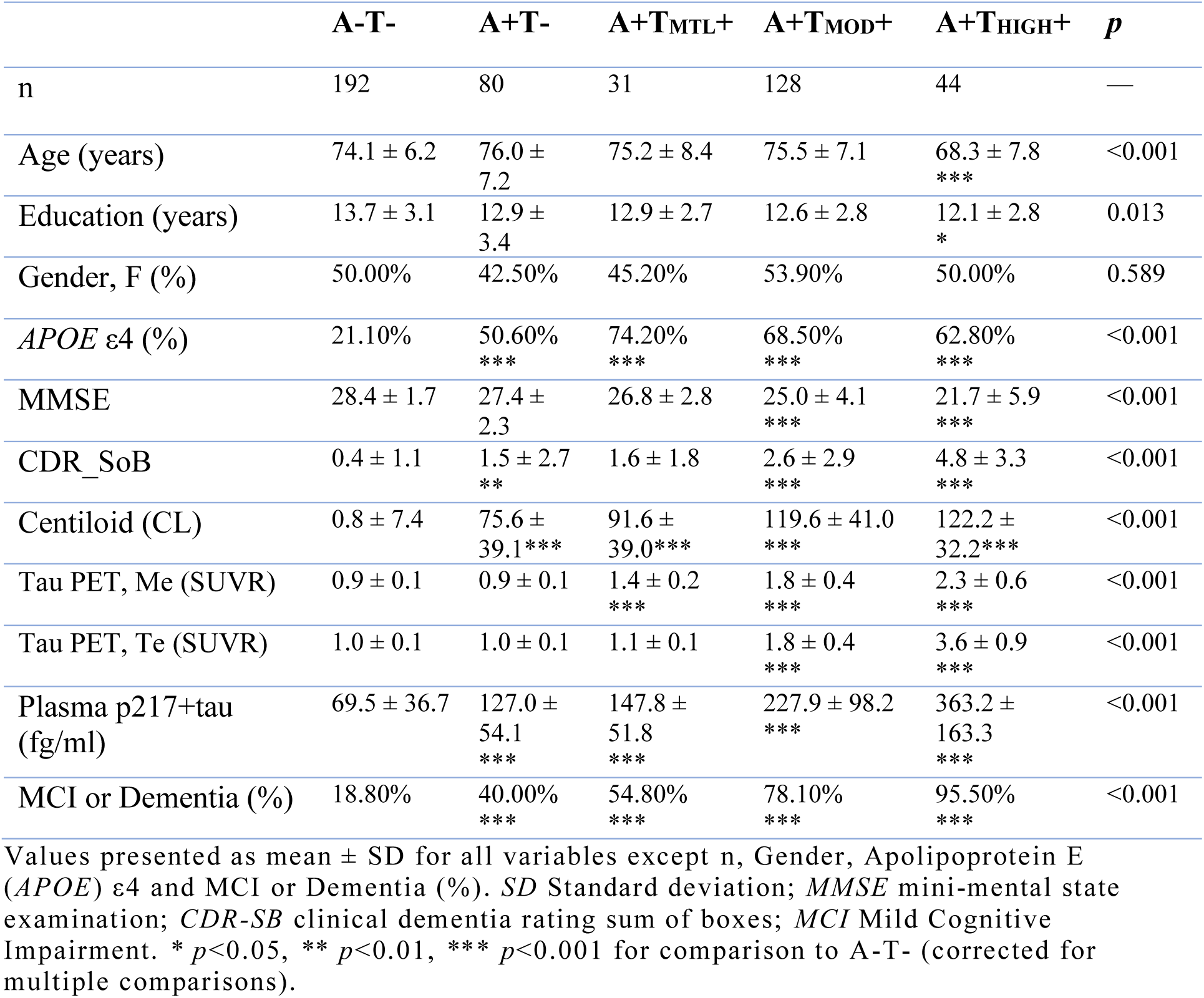
Demographic characteristics.

### Plasma p217+tau concentration by PET stages

The median concentration of plasma p217+tau exhibited a clear incremental trend across the stages, with values of 63.5 fg/ml in the A-T-, 118.7 fg/ml in A+T-, 140.7 fg/ml in A+T_MTL_+, 211.0 fg/ml in A+T_MOD_+ and 336.7 fg/ml in A+T_HIGH_+ (Figure 1). A pairwise comparison— corrected for multiple comparisons— revealed significant differences in mean plasma p217+tau concentration between all stages, except for the comparison between A+T- and A+T_MTL_+. Henceforth, in all subsequent analyses, these two stages were consolidated into a unified stage referred to as Initial/Early (A+T_None/MTL_). For detailed information on Cohen’s *d* effect sizes and *p*-values for comparison between the stages, see Supplementary Table 3.

**Figure 1.**
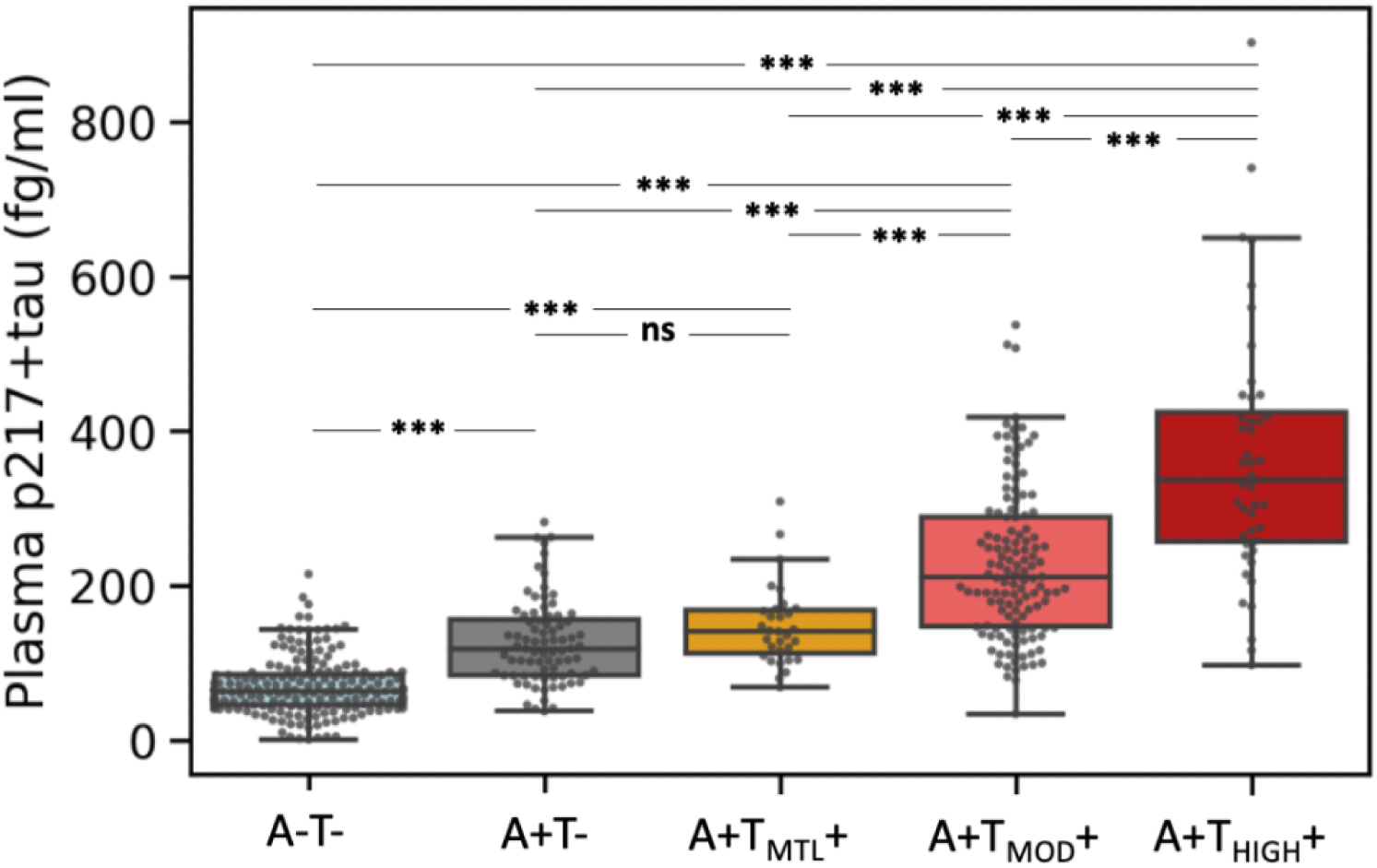
Plasma p217+tau levels in the different PET stages. The error bars in the boxplots represent 1.5 times the interquartile range (IQR) above and below the upper (75 ^th^ percentile) and lower (25 ^th^ percentile) quartiles. Plasma p217+tau concentrations exhibited a clear incremental trend across these stages, with significant differences between all, except for the Initial and Early stages. *A-T-* amyloid negative and tau negative; *A+T-* amyloid positive and tau negative; *A+T_MTL_+* amyloid positive with tau uptake limited to medial temporal region; *A+T_MOD_+* amyloid positive with moderate tau uptake in temporo-parietal region; *A+T_HIGH_+* amyloid positive with high tau uptake in temporo-parietal region. *\**\*\* *p*<0.001 (corrected for multiple comparisons); *ns* non-significant.

### Group-level approach for disease staging

#### Discrimination between A-T- and A+ stages

In a clinical trial investigating the effect of a disease-modifying treatment on all biological PET stages of AD, discriminating between those stages and A-T- would be important. Our findings showed that when CU and CI participants were combined, plasma p217+tau alone discriminated between A-T- and all stages combined of A+, with an AUC of 0.92 (Figure 2A), significantly better than the discrimination by the base model which included age, gender and *APOE* ε4 status (AUC: 0.75, *p*<0.001). The Youden index provided a threshold concentration of 99.4 fg/ml [89.7 - 102.4 fg/ml], which yielded sensitivity of 0.87 [0.83 - 0.91], specificity of 0.84 [0.78 - 0.88], PPV of 0.89 [0.85 - 0.92] and NPV of 0.81 [0.77 - 0.87] (Supplementary Table 4). Adding age or gender separately did not improve the AUC of the model with p217+tau while *APOE* ε4 status improved the model slightly (AUC: 0.94, *p*<0.05). The full model including p217+tau, age, gender and *APOE* ε4 status yielded the highest AUC (0.94, *p*<0.01 compared to model with p217+tau only).

**Figure 2.**
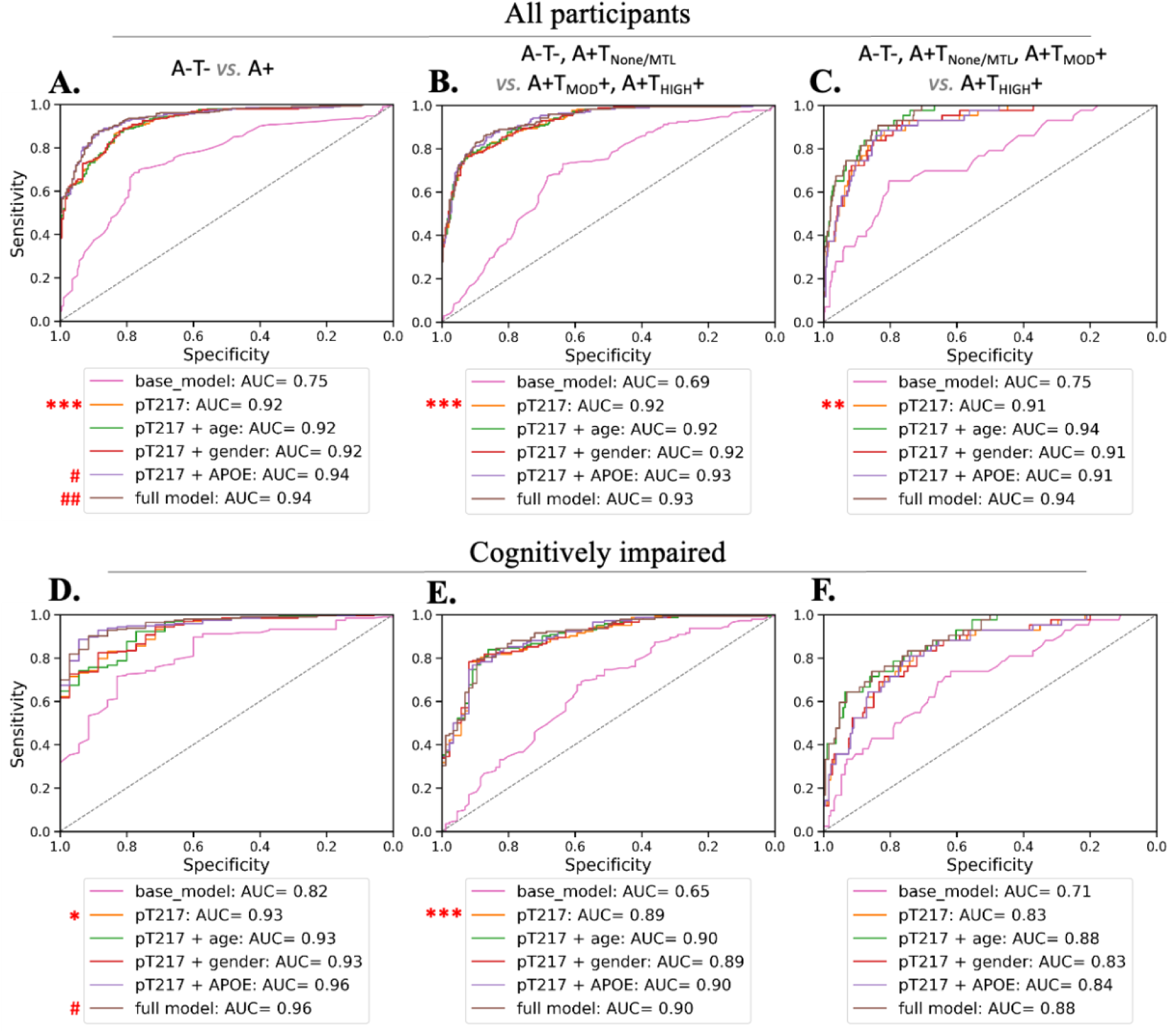
ROC analysis for group-level PET staging, CU and CI combined (Top panel), and CI only (Bottom panel). Base model includes age, gender, and Apolipoprotein E (*APOE*) ε4 status. Full model includes p217+tau, age, gender, and *APOE* ε4 status. *pT217* plasma p217+tau; *AUC* area under the receiver operating characteristic curve. *A-T-* amyloid negative and tau negative; *A+T_None/MTL_* amyloid positive and tau negative or amyloid positive with tau uptake limited to medial temporal region; *A+T_MOD_+* amyloid positive with moderate tau uptake in temporo-parietal region; *A+T_HIGH_+* amyloid positive with high tau uptake in temporo-parietal region. *\*p*<0.05, *\**\**p*<0.01, *\**\*\**p*<0.001 for comparison of the model with p217+tau only, to the base model (corrected for multiple comparisons). *^#^ p*<0.05, *^##^ p*<0.01, for comparison of models with additional predictors to the model with p217+tau only (after correction for multiple comparisons).

Among CI only participants, plasma p217+tau discriminated between A-T- and A+ individuals with an AUC of 0.93 (Figure 2D), significantly better than the discrimination by the base model (AUC: 0.82, *p*<0.05). Youden threshold of 126.7 fg/ml [94.8 - 149.9 fg/ml] gave a sensitivity of 0.83 [0.69 - 0.95], specificity of 0.86 [0.79 - 1.0], PPV of 0.97 [0.96 - 1.0], and NPV of 0.48 [0.34 - 0.74] (Supplementary Table 5). Adding age, gender or *APOE* ε4 status separately did not improve the model with p217+tau. However, adding all predictors together (full model) had the highest AUC (0.96, *p*<0.05 compared to model with p217+tau only).

#### Discrimination between combined A-T-/A+T_None/MTL_ and combined A+T_MOD_+/A+T_HIGH_+

In a clinical trial assessing the impact of a disease-modifying treatment on individuals with Aβ and intermediate or high neocortical tau pathology, screening for these individuals *vs.* A- T-/A+T_None/MTL_ individuals becomes a crucial consideration.

Among the CU & CI combined, for discriminating these two groups, using p217+tau alone, the AUC was 0.92 (Figure 2B), performing significantly better than the base model (AUC: 0.69, *p*<0.001).Youden index gave a threshold of 168.0 fg/ml [131.8 - 177.4 fg/ml] which yielded a sensitivity of 0.77 [0.73 - 0.90], specificity of 0.91 [0.80 - 0.95], PPV of 0.84 [0.71- 0.89], and NPV of 0.88 [0.85 - 0.93] (Supplementary Table 4). Adding age, gender or *APOE* ε4 status separately or together did not improve the AUC of the model with p217+tau. Among the CI only, the AUC was 0.89 (Figure 2E), significantly better than the discrimination by the base model (AUC: 0.65, *p*<0.001), Youden threshold 177.4 fg/ml [168.0 - 179.4 fg/ml], sensitivity 0.78 [0.74 - 0.86], specificity 0.92 [0.85 - 0.96], PPV 0.94 [0.89 - 0.97], NPV 0.72 [0.66 - 0.81] (Supplementary Table 5). Adding age, gender or *APOE* ε4 status separately or together did not improve the model with p217+tau.

#### Discrimination between combined A-T-/A+T_None/MTL_/A+T_MOD_+ and A+T_HIGH_+

In a therapeutic trial evaluating the effect of an intervention in the Advanced (high neocortical tau) group only, or wishing to exclude those with Advanced tau, discrimination between A-T-/A+T_None/MTL_/A+T_MOD_+ and A+T_HIGH_+ will be crucial.

Among the CU & CI combined, the use of plasma p217+tau alone for discriminating between these two groups gave an AUC of 0.91 (Figure 2C), significantly better than the discriminatory performance of the base model (AUC: 0.75, *p*<0.01). Youden index yielded a threshold of 205.4 fg/ml [173.2 - 257.9 fg/ml] with a sensitivity of 0.89 [0.79 - 0.97], specificity of 0.82 [0.75 - 0.90], PPV of 0.33 [0.25 - 0.47], NPV of 0.99 [0.98 - 1.0] (Supplementary Table 4). Adding age, gender or *APOE* ε4 status separately or together did not improve the model with p217+tau.

When only CI participants were included, plasma p217+tau alone discriminated between these two groups with an AUC of 0.84 (Figure 2F) and this AUC did not significantly differ from any of the other models. Youden index provided a threshold of 230.3 fg/ml [205.4 - 299.5 fg/ml] giving sensitivity of 0.84 [0.69 - 0.95], and specificity 0.71 [0.64 - 0.88] (Supplementary Table 5). With PPV of 0.39 [0.31 - 0.58] and NPV of 0.95 [0.92 - 0.98], p217+tau effectively excluded Advanced stage AD when less than the Youden threshold but did not accurately predict Advanced stage when above, predominantly due to the presence of many Intermediate stage AD participants with p217+tau above the threshold.

For a pairwise comparison of each two biological PET stages, see Supplementary Table 6.

#### Discrimination between disease stages of enrolled participants

In the preceding sections, we demonstrated that employing a Youden threshold of 177.4 fg/ml allowed screening for CI participants who were either A+T_MOD_+ or A+T_HIGH_+, with a PPV of 0.94. In some therapeutic trials, it might be essential to stage enrolled participants for analytical purposes. For instance, one may wish to assess the impact of an intervention only in participants with high neocortical tau (A+T_HIGH_+). This led us to enquire whether, in the absence of Aβ and tau PET imaging, p217+tau could discriminate between A+T_HIGH_+ and those at lower stages in participants with p217+tau values above 177.4 fg/ml (pT_177.4_+).

Among the pT_177.4_+ participants, for discriminating between the A+T_HIGH_+ and the lower stages, the AUC was 0.76 [0.68 - 0.83], Youden index threshold 299.5 fg/ml [257.9 - 402.9], sensitivity 0.71 [0.47 - 0.91], specificity 0.70 [0.52 - 0.95], PPV 0.52 [0.41 - 0.79], NPV 0.84 [0.76 - 0.93]. This indicates that this threshold could be potentially used to rule out A+T_HIGH_+, among the pT_177.4_+ participants.

### Individual-level approach for disease staging

We also performed logistic regressions to ascertain whether an individual’s plasma p217+tau level can inform their classification into A-T-, Initial-Early (A+T_None/MTL_), Intermediate (A+T_MOD_+), or Advanced (A+T_HIGH_+) stages. Figure 3 (left: CU & CI combined; right: CI only) displays the logistic regression probability scores for each stage as a function of plasma p217+tau concentrations. At any given plasma p217+tau value, an individual is most likely to belong to the stage with the highest probability score. Individuals with low p217+tau concentrations (below 110 fg/ml for CU & CI combined, or 93 fg/ml for CI only) had the highest likelihood of being A-T-. A+T_None/MTL_ was the classification with the highest likelihood in p217+tau value range of 110-172 fg/ml for CU & CI combined, and 93-150 fg/ml for CI only. A+T_MOD_+ was the most likely classification for a wide range of moderate p217+tau values (172-375 fg/ml for CU & CI combined, and 150-374 fg/ml for CI only). For p217+tau values higher than ∼375 fg/ml, the classification of A+T_HIGH_+ was the most likely. In Figure 3, the likelihoods from logistic regression (solid line) are compared to the ones estimated using the sliding window method (dots), indicating good concordance between the two approaches in computing probabilities.

**Figure 3.**
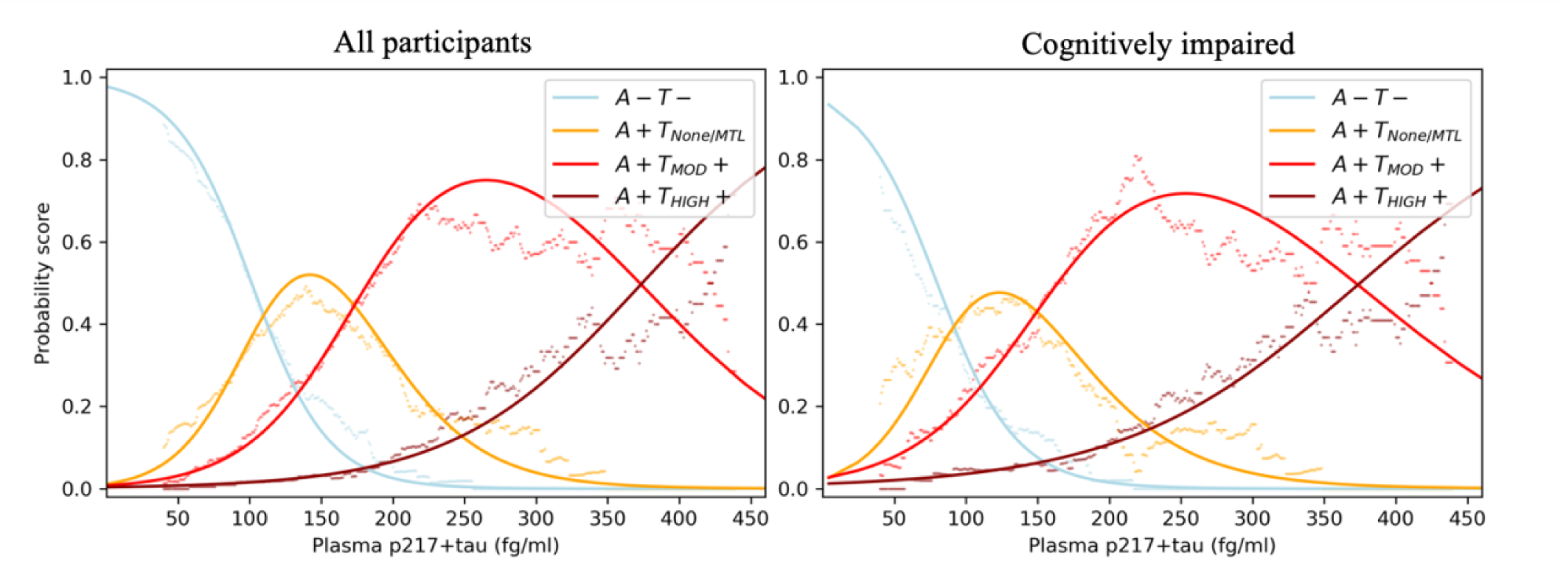
Logistic regression probability scores *vs*. plasma p217+tau concentration values. CU & CI combined, on the left. CI only on the right. Line is the probability scores generated by logistic regression and dots are the probability scores generated by sliding window methods. *A-T-* amyloid negative and tau negative; *A+T_None/MTL_* amyloid positive and tau negative or amyloid positive with tau uptake limited to medial temporal region; *A+T_MOD_+* amyloid positive with moderate tau uptake in temporo-parietal region; *A+T_HIGH_+* amyloid positive with high tau uptake in temporo-parietal region.

## Discussion

In the present study, we evaluated the ability of plasma p217+tau to discriminate between AD disease stages, employing the biological PET stages recommended by draft NIA-AA Revised Criteria 2023 ^9^ where we have defined the Advanced stage as >80 Centaur (2.68 SUVR_temporo-parietal_). We showed that as disease progressed from A-T- to the Advanced biological PET stages (A+T_HIGH_+), there was a clear incremental trend in plasma p217+tau concentration, with A+T- or A+T_MTL_+ experiencing an almost two-fold increase, A+T_MOD_+ more than a three-fold increase, and A+T_HIGH_+ more than a five-fold increase in p217+tau, relative to A- T-.

Next, our group-level analysis was performed with a clinical trial in mind, where establishing cut-off points allowed screening individuals at certain disease stages in or out of the trial or enabled grouping enrolled participants for analytical purposes. This is critical as clinical trials move to recruit participants based on not merely Aβ or tau positive or negative status, but enroll based on magnitude and topographical distribution of Aβ and tau (such as TRAILBLAZER-ALZ 2 ^1^). With high cost and limited availability of PET, predicting level and spatiotemporal patterns of AD pathology, particularly for tau, using plasma biomarkers becomes a priority.

Group-level examination of how well plasma p-tau217 can distinguish between different Braak-like PET stages in CU has been reported in a recent study by Jack *et al*. ^23^. They found that p-tau217 discriminated between tau PET negative and tau PET positive (any Braak-like PET stage) with an AUC of 0.83 which was not significantly different from the discrimination by Quanterix p-tau181 or a base model including age, sex and *APOE*. Only 37% of our CU were A+, and among these, only a few had high neocortical tau burden, so our study lacked the statistical power to discriminate between AD biological PET stages within the CU group. Thus, we chose to limit our report to either the CU & CI combined group or CI-only group.

Here, we showed that if screening for participants with early symptomatic AD, with Aβ and intermediate/high neocortical tau (similar to TRAILBLAZER-ALZ 2), plasma p217+tau could discriminate between participants at these stages and other CI participants with an AUC of 0.89, and if the Youden threshold derived in this cohort for this purpose of 177.4 fg/ml was used, 94% of p217+tau positive participants would be in the target PET stages (Intermediate/Advanced: A+T_MOD_+ or A+T_HIGH_+). However, when investigating the utility of plasma p217+tau for separating advanced (A+T_HIGH_+) stage of AD in the participants above the p217+tau threshold of 177.4 fg/ml, the AUC was lower (0.75) and the Youden threshold gave a NPV of 0.84 with a PPV of only 0.51. Tau PET scan would still be essential for separating A+T_MOD_+ from A+T_HIGH_+.

Alternatively, if a trial were to enrol symptomatic participants but exclude those with Advanced stage AD (A+T_HIGH_+), we showed that 95% of CI participants with plasma p217+tau < 230.3 fg/ml were in disease stages lower than A+T_HIGH_+ and therefore this threshold could be appropriate for exclusion of A+T_HIGH_+.

The pairwise discrimination of each pair of AD biological PET stages —Initial *vs*. Early, Early *vs*. Intermediate, and Intermediate *vs*. High— yielded lower AUC values. The AUC of 0.64 for Initial *vs.* Early, suggested that p217+tau did not have adequate discriminatory performance for these two stages and provided additional support for combining these two into one stage for subsequent analyses. The AUC of 0.76, observed in the discrimination between Early and Intermediate stages, falls within an acceptable yet relatively lower range. This highlights that the higher AUC of 0.92 reported for the A-T-/Initial/Early *vs.* Intermediate/Advanced discrimination is predominantly influenced by the inclusion of more extreme values from A-T- and Advanced stages in the discrimination. The same reasoning applies to the AUC of 0.77 for Intermediate *vs.* High relative to the AUC of 0.91 for A-T-/Initial/Early*/*Intermediate *vs.* Advanced.

Previous studies have shown that ^18^F-MK6240 might be better suited to detect early tau accumulation (Me ROI) compared to ^18^F-Flortaucipir ^24, 25^, thus our use of ^18^F-MK6240 as tau PET tracer (as compared to Flortaucipir in the study by Jack *et al*.) may have provided a more optimal identification of Early stage AD. Yet our findings with ^18^F-MK6240 were not able to show that plasma p217+tau was able to discriminate A+T- from A+T_MTL_+.

In a patient-centric approach, an individual’s absolute risk of having AD pathology at a given plasma biomarker concentration may be more relevant for clinical decision-making. Therefore, we developed a risk prediction model using p217+tau as the predictor and estimated probability scores —which are individuals’ absolute risk— of falling within a disease stage for their individual p217+tau level. The plot of the likelihood of PET stage *vs*. p217+tau concentration showed good discrimination of A-T- vs any A+ stage and of combined Intermediate/Advanced disease stage vs lower stages in the CI. However, with the overlap in likelihood functions for most PET stages, confident staging of an individual with plasma p217+tau alone is limited. One instead should consider an individual participant’s likelihood for belonging to each of the stages based on their p217+tau level as illustrated by Figure 3.

A limitation of this study is that the findings are derived from a research cohort with a relatively high prevalence of Aβ-PET positivity in the three clinical groups. Replication of prevalence-dependent findings (i.e., PPV, NPV, estimated probability scores) in a “real-world” memory clinic setting is needed before any of the findings could be applied in clinical practice. Moreover, future studies are needed to develop risk prediction models using a combination of several factors to accurately predict individual-level disease stages. To conclude, plasma p217+tau performs well for group-level discrimination between disease stages (except Initial and Early), a useful property for clinical trial screening and enrichment.

## Data Availability

All data produced in the present study are available upon reasonable request to the authors, after obtaining approval from AIBL and ADNeT.

## Acknowledgments

Some data used in this research article were obtained from the Australian Imaging Biomarkers and Lifestyle flagship study of aging (AIBL), funded by the Commonwealth Scientific and Industrial Research Organization (CSIRO), National Health and Medical Research Council (NHMRC), and other participating institutions. Some data were obtained from the Australian Dementia Network (ADNeT) that receives funding support from the NHMRC. AIBL researchers are listed on the website www.aibl.csiro.au. The authors thank those who participated in the study, as well as their families.

## Funding

The research was supported by the Australian Federal Government through NHMRC grants APP1132604, APP1140853 and APP1152623 and by a grant from Enigma Australia. Janssen Pharmaceuticals paid a commercial data access fee to the AIBL study of ageing. Janssen Pharmaceuticals performed the p217+tau measurements but did not perform the data analysis. The co-authors employed by Janssen did provide comments on the manuscript and Janssen provided approval for submission of the manuscript. The other sponsors had no role in the design and conduct of the study, in the collection, analysis, and interpretation of data, or in the preparation of the manuscript.

## Competing interests

Christopher C. Rowe has received research grants from NHMRC, Enigma Australia, Biogen, Eisai and Abbvie. He is on the scientific advisory board for Cerveau Technologies and has consulted for Prothena, Eisai, Roche, and Biogen Australia. Victor L. Villemagne is and has been a consultant or paid speaker at sponsored conference sessions for Eli Lilly, Life Molecular Imaging, GE Healthcare, IXICO, Abbvie, Lundbeck, Shanghai Green Valley Pharmaceutical Co Ltd, AC Immune and Hoffmann La Roche. Ziad S. Saad, Gallen Triana-Baltzer, and Hartmuth C. Kolb are employees of Janssen Pharmaceuticals. Simon M. Laws is a scientific advisor for Cytox Ltd. The other authors did not report any conflict of interest.

## Notes

### Funding Statement

The study was supported by the Australian Federal Government through NHMRC grants APP1132604, APP1140853 and APP1152623 and by a grant from Enigma Australia. Janssen Pharmaceuticals paid a commercial data access fee to the AIBL study of ageing.

### Author Declarations

Institutional ethics review committees of the AIBL and ADNeT studies gave ethical approval for this work.

