## Supplementary Information for "Alzheimer’s Disease biological PET staging using plasma p217+tau"

Supplementary Table 1. Demographic characteristics of participants that did not fit the NIA-AA based profiles or the A-T- group

| | **A-$T_{Me+Te-R-}$** | **A-$T_{Me-Te+R-}$** | **A-$T_{Me-Te-R+}$** | **A-$T_{Me-Te+R+}$** | **A-$T_{Me+Te+R-}$** | **A-$T_{Me+Te+R+}$** |
|---|---|---|---|---|---|---|
| **n** | 11 | 2 | 6 | 3 | 4 | 5 |
| **Age (years)** | 76.4 ± 5.8 | 79.0 ± 3.0 | 74.7 ± 5.2 | 67.0 ± 5.0 | 78.0 ± 4.1 | 70.2 ± 4.3 |
| **Education (years)** | 13.5 ± 4.3 | 12.0 ± 3.0 | 15.0 ± 2.2 | 16.0 ± 4.5 | 14.0 ± 1.7 | 11.0 ± 2.8 |
| **Gender, F (%)** | 45% | 50% | 83% | 67% | 7% | 40% |
| **MMSE** | 26.4 ± 3.6 | 29.5 ± 0.5 | 29.2 ± 0.7 | 27.3 ± 2.1 | 25.5 ± 3.4 | 24.0 ± 2.6 |
| **CDR_ SoB** | 1.4 ± 2.0 | 0.2 ± 0.2 | 0.1 ± 0.2 | 2.5 ± 2.5 | 1.4 ± 1.0 | 3.0 ± 1.7 |
| **Tau PET SUVR (MetaT)** | 1.2 ± 0.1 | 1.1 ± 0.1 | 1.0 ± 0.1 | 1.1 ± 0.0 | 1.3 ± 0.2 | 1.8 ± 0.9 |
| **Centiloid (CL)** | 5.6 ± 8.0 | -6.2 ± 1.2 | -4.1 ± 5.1 | -3.1 ± 3.4 | -0.6 ± 9.2 | 9.2 ± 7.6 |
| **Plasma p217+tau (fg/ml)** | 85.8 ± 48.2 | 126.2 ± 57.9 | 57.8 ± 25.5 | 63.0 ± 30.0 | 112.8 ± 31.2 | 159.3 ± 94.7 |
| **MCI or Dementia (%)** | 45% | 0% | 17% | 67% | 75% | 80% |

Values presented as mean ± SD for all variables except n, Gender, Apolipoprotein E (*APOE*) ε4 and MCI or Dementia (%). *MMSE* mini-mental state examination; *CDR-SoB* clinical dementia rating sum of boxes; *SD* Standard deviation; *Me* mesial temporal ROI; *Te* temporoparietal ROI; *R* rest of neocortex. Positive sign denotes being above the pre-defined thresholds and negative below the pre-defined thresholds outlined in the Methods section. *Tau PET SUVR (*MetaT*)* Tau $^{18}$F-MK6240 SUVR estimated in a standard meta-temporal composite region, comprising entorhinal cortex, parahippocampus, amygdala, inferior temporal, fusiform, and middle temporal cortex ROI.

Supplementary Table 2. Demographic characteristics, breakdown by clinical diagnosis

| | | **CU** | **MCI** | **Dementia** |
|---|---|---|---|---|
| n | | 248 | 144 | 83 |
| Age (years) | | 75.6 ± 6.2 | 73.8 ± 7.7 | 71.5 ± 8.2 |
| Education (years) | | 13.7 ± 3.0 | 12.6 ± 3.1 | 12.2 ± 2.9 |
| Gender, Female (%) | | 54% | 45% | 43% |
| *APOE* ε4 (%) | | 30% | 40% | 55% |
| MMSE | | 28.6 ± 1.4 | 26.4 ± 2.6 | 22.2 ± 5.0 |
| CDR_SoB | | 0.2 ± 0.5 | 1.2 ± 0.8 | 5.6 ± 3.0 |
| Centiloid (CL) | | 32.9 ± 50.6 | 90.2 ± 56.1 | 103.5 ± 52.6 |
| Aβ+ PET (%) | | 37% | 81% | 90% |
| Tau PET, meta temporal (SUVR) | | 1.0 ± 0.2 | 1.6 ± 0.6 | 2.0 ± 0.8 |
| Plasma p217+tau (fg/ml) | | 99.3 ± 76.4 | 197.4 ± 123.3 | 243.1 ± 144.4 |
| AT biomarker groups | A-T- (%) | 63% | 19% | 10% |
| | A+T- (%) | 19% | 16% | 11% |
| | $A+T_{MTL}+$ (%) | 6% | 7% | 8% |
| | $A+T_{MOD}+$ (%) | 11% | 49% | 36% |
| | $A+T_{HIGH}+$ (%) | 1% | 9% | 35% |

Values presented as mean ± SD for all variables except n, Gender, Apolipoprotein E (*APOE*) ε4, Aβ+ PET% and AT biomarker groups. Total n = 475. Aβ+ PET based on a threshold of 25 CL. *MMSE* mini-mental state examination; *CDR-SoB* clinical dementia rating sum of boxes; *SD* Standard deviation; *CU* Cognitively Unimpaired; *MCI* Mild Cognitive Impairment; *A-T-* amyloid negative and tau negative; *A+T-* amyloid positive and tau negative; $A+T_{MTL}+$ amyloid positive with tau uptake limited to medial temporal region; $A+T_{MOD}+$ amyloid positive with moderate tau uptake in temporo-parietal region; $A+T_{HIGH}+$ amyloid positive with high tau uptake in temporo-parietal region.

Supplementary Table 3. The *p*-values and Cohen's *d* effect sizes for differences in plasma p217+tau concentration

| **Comparisons** | **Adjusted *p* values** | **Cohen's *d*** |
|---|---|---|
| A-T- *vs*. A+T- | <0.001 | 1.3 |
| A-T- *vs*. A+T$_{MTL}$+ | <0.001 | 2.0 |
| A-T- *vs*. A+T$_{MOD}$+ | <0.001 | 2.3 |
| A-T- *vs*. A+T$_{HIGH}$+ | <0.001 | 3.8 |
| A+T- *vs*. A+T$_{MTL}$+ | 0.731 | 0.4 |
| A+T- *vs*. A+T$_{MOD}$+ | <0.001 | 1.2 |
| A+T- *vs*. A+T$_{HIGH}$+ | <0.001 | 2.2 |
| A+T$_{MTL}$+ *vs*. A+T$_{MOD}$+ | <0.001 | 0.9 |
| A+T$_{MTL}$+ *vs*. A+T$_{HIGH}$+ | <0.001 | 1.6 |
| A+T$_{MOD}$+ *vs*. A+T$_{HIGH}$+ | <0.001 | 1.1 |

Multiple comparison of group means, using Tukey HSD. *A-T-* amyloid negative and tau negative; *A+T-* amyloid positive and tau negative; *A+*$T_{MTL}$+ amyloid positive with tau uptake limited to medial temporal region; *A+*$T_{MOD}$+ amyloid positive with moderate tau uptake in temporo-parietal region; *A+*$T_{HIGH}$+ amyloid positive with high tau uptake in temporo-parietal region.

Supplementary Table 4. Statistics of the ROC analysis to predict disease stage using p217+tau Youden threshold, *CU and CI participants combined*

| | AUC | Youden threshold | Sensitivity | Specificity | PPV | NPV |
|---|---|---|---|---|---|---|
| Model 1 | 0.92 [0.90 - 0.94] | 99.38 [89.75 - 102.45] | 0.87 [0.83 - 0.91] | 0.84 [0.78 - 0.88] | 0.89 [0.85 - 0.92] | 0.81 [0.77 - 0.87] |
| Model 2 | 0.92 [0.90 - 0.94] | 168.04 [131.83 - 177.38] | 0.77 [0.73 - 0.90] | 0.91 [0.80 - 0.95] | 0.84 [0.71 - 0.89] | 0.88 [0.85 - 0.93] |
| Model 3 | 0.91 [0.88 - 0.95] | 205.43 [173.18 - 257.86] | 0.89 [0.79 - 0.97] | 0.82 [0.75 - 0.9] | 0.33 [0.25 - 0.47] | 0.99 [0.98 - 1.0] |

Values reported with bootstrapped 95% confidence intervals, shown in square brackets. Model 1: A-T- vs. A+T-/A+$T_{MTL}$+/A+$T_{MOD}$+/A+$T_{HIGH}$+. The reported AUC value was slightly higher and Youden's index threshold lower than previously reported for this p-tau assay when discriminating based on Aβ status (which was based on cohort 1 p217+tau data) [4]. This is attributed to the cohort 2 differences due to between-lab variations (for details on cohort 1 and cohort 2, see Methods). It is less likely to be due to exclusion of A-T+ individuals from the current study, as repetition of this analysis, with the 31 A-T+ included, yielded an AUC of 0.91 [0.89 - 0.93] and Youden threshold of 99.38 fg/ml [89.75 - 126.68]. PPV and NPV reported at A+ observed prevalence of 58%.

Model 2: A-T-/A+T-/A+$T_{MTL}$+ vs. A+$T_{MOD}$+/A+$T_{HIGH}$+. PPV and NPV reported at A+$T_{MOD}$+/A+$T_{HIGH}$+ observed prevalence of 33%.

Model 3: A-T-/A+T-/A+$T_{MTL}$+/A+$T_{MOD}$+ vs. A+$T_{HIGH}$+. PPV and NPV reported at A+$T_{HIGH}$+ observed prevalence of 24%.

*A-T-* amyloid negative and tau negative; *A+T-* amyloid positive and tau negative; *A+$T_{MTL}$+* amyloid positive with tau uptake limited to medial temporal region; *A+$T_{MOD}$+* amyloid positive with moderate tau uptake in temporo-parietal region; *A+$T_{HIGH}$+* amyloid positive with high tau uptake in temporo-parietal region; *PPV* positive predictive value; *NPV* negative predictive value.

Supplementary Table 5. Statistics of the ROC analysis to predict disease stage using p217+tau Youden threshold, *in CI participants*

| | AUC | Youden threshold | Sensitivity | Specificity | PPV | NPV |
|---|---|---|---|---|---|---|
| Model 1 | 0.93 [0.89 - 0.96] | 126.68 [94.83 - 149.88] | 0.83 [0.69 - 0.95] | 0.86 [0.79 - 1.0] | 0.97 [0.96 - 1.0] | 0.48 [0.34 - 0.74] |
| Model 2 | 0.89 [0.86 - 0.93] | 177.38 [168.04 - 179.39] | 0.78 [0.74 - 0.86] | 0.92 [0.85 - 0.96] | 0.94 [0.89 - 0.97] | 0.72 [0.66 - 0.81] |
| Model 3 | 0.84 [0.78 - 0.89] | 230.29 [205.43 - 299.52] | 0.84 [0.69 - 0.95] | 0.71 [0.64 - 0.88] | 0.39 [0.31 - 0.58] | 0.95 [0.92 - 0.98] |

Values reported with bootstrapped 95% confidence intervals, shown in square brackets.
Model 1: A-T- *vs.* A+T-/A+$T_{MTL}$+/A+$T_{MOD}$+/A+$T_{HIGH}$+. PPV and NPV reported at A+ observed prevalence of 72%.

Model 2: A-T-/A+T-/A+$T_{MTL}$+ *vs.* A+$T_{MOD}$+/ A+$T_{HIGH}$+. PPV and NPV reported at A+$T_{MOD}$+/A+$T_{HIGH}$+ observed prevalence of 51%.

Model 3: A-T-/A+T-/A+$T_{MTL}$+/A+$T_{MOD}$+ *vs.* A+$T_{HIGH}$+. PPV and NPV reported at A+$T_{HIGH}$+ observed prevalence of 39%.

*A-T-* amyloid negative and tau negative; *A+T-* amyloid positive and tau negative; *A+$T_{MTL}$+* amyloid positive with tau uptake limited to medial temporal region; *A+$T_{MOD}$+* amyloid positive with moderate tau uptake in temporo-parietal region; *A+$T_{HIGH}$+* amyloid positive with high tau uptake in temporo-parietal region; *PPV* positive predictive value; *NPV* negative predictive value.

Supplementary Table 6. Discrimination between only two stages at a time, *CU and CI participants combined*

| | AUC | Youden threshold | Sensitivity | Specificity | PPV | NPV |
|---|---|---|---|---|---|---|
| A+T- *vs.* A+$T_{MTL}$+ | 0.57 [0.43 - 0.71] | 164.10 [80.17 - 266.58] | 0.41 [0.25 - 1.00] | 0.79 [0.16 - 0.97] | 0.46 [0.22 - 0.75] | 0.73 [0.65 - 1.0] |
| A+$T_{MTL}$+ *vs.* A+$T_{MOD}$+ | 0.78 [0.68 - 0.87] | 178.91 [144.24 - 179.98] | 0.73 [0.67 - 0.87] | 0.82 [0.62 - 0.95] | 0.96 [0.92 - 0.99] | 0.34 [0.24 - 0.51] |
| A+$T_{MOD}$+ *vs.* A+$T_{HIGH}$+ | 0.74 [0.67 - 0.82] | 299.52 [230.29 - 358.75] | 0.63 [0.51 - 0.89] | 0.77 [0.53 - 0.9] | 0.53 [0.39 - 0.7] | 0.83 [0.79 - 0.93] |

To discriminate between only two biological PET stages at a time, the sample sizes were smaller, thus we performed this analysis only on the combined CU and CI participants. The AUC values were expectedly lower when using plasma p217+tau to discriminate between only two stages at a time. Values reported with bootstrapped 95% confidence intervals, shown in square brackets. *A+T-* amyloid positive and tau negative; $A+T_{MTL}+$ amyloid positive with tau uptake limited to medial temporal region; $A+T_{MOD}+$ amyloid positive with moderate tau uptake in temporo-parietal region; $A+T_{HIGH}+$ amyloid positive with high tau uptake in temporo-parietal region.
